# Retinal transcriptome and cellular landscape in relation to the progression of diabetic retinopathy

**DOI:** 10.1101/2022.05.05.22274746

**Authors:** Jiang-Hui Wang, Raymond C.B. Wong, Guei-Sheung Liu

## Abstract

**Importance:** Previous studies identify putative genes associated with diabetic retinopathy only focusing on specific clinical stage, thus resulting genes are not necessarily reflective of disease progression.

**Objective:** To identify genes associated with the severity level of diabetic retinopathy using likelihood-ration test (LRT) and ordinal logistic regression (OLR) model, as well as to profile immune and retinal cell landscape in progressive diabetic retinopathy using a machine learning deconvolution approach.

**Design, Setting, and Participants:** This study used published transcriptomic dataset (GSE160306) from macular regions of donors with different degrees of diabetic retinopathy (10 healthy controls, 10 cases of diabetes, 9 cases of nonproliferative diabetic retinopathy and 10 cases of proliferative diabetic retinopathy or combined with diabetic macular edema). Transcriptomic dataset was released on April 19, 2021; data were analyzed on 28th March 2022.

**Main Outcomes and Measures:** Identification of severity-associated genes with LRT and OLR model and proportional changes of immune and retinal cells in progressive diabetic retinopathy.

**Results:** By controlling for gender and age using LRT and OLR, 50 genes were identified to be significantly increased in expression with the severity of diabetic retinopathy. Functional enrichment analyses suggested these severity-associated genes are related to inflammation and immune responses. *CCND1* and *FCGR2B* are further identified as key regulators to interact with many other severity-associated genes and are crucial to inflammation. Deconvolution analyses demonstrated that the proportions of memory B cells, M2 macrophages and Müller glia were significantly increased with the progression of diabetic retinopathy.

**Conclusions and Relevance:** These findings demonstrate the deep analyses of transcriptomic data can advance our understanding of progressive ocular diseases, such as DR, by applying LRT and OLR model as well as bulk gene expression deconvolution.

**Key Points:** *Question:* Do genes change in expression or immune cells and retinal cells change in proportions in the human retina with the severity of diabetic retinopathy?

*Findings:* In this in-depth retinal transcriptomic analysis of 39 participants, consisting of 10 heathy controls, 29 cases of diabetic retinopathy at different clinical stages, severity-associated genes were identified, and the estimated proportions of immune cell and retinal cells were profiled, respectively.

*Meaning:* These findings demonstrate that a group of genes are significantly increased in expression, and the proportions of memory B cells, M2 macrophages and Müller glia were significantly increased with the severity of diabetic retinopathy.

## Introduction

Diabetic retinopathy (DR) is a progressive retinal complication of diabetes, causing significant visual impairment on a global scale.^1^ Microvascular lesions of retina have been used as the manifestation to evaluate and classify the severity level of DR. There are 2 broad categories, including the early stage of nonproliferative diabetic retinopathy (NPDR) and the advanced stage of proliferative diabetic retinopathy (PDR).^1^ NPDR is manifested by increased vascular permeability and capillary occlusion, while PDR is characterised by retinal neovascularization.^2^ An important addition category of DR is diabetic macular edema (DME), the most common cause of vision loss in patients with DR.^2^ Genetic studies including candidate gene studies, linkage studies, powerful genome-wide association studies (GWAS) and meta-analysis have identified putative candidate gene or single nucleotide polymorphisms associated with DR.^3,4^ While these studies provide critical information that helps to interpret the mechanisms of DR, they mostly study one specific type of DR thus the resulting putative genes are not likely to be associated with DR progression.

In this study, we assess the macular transcriptomic data from donors with different severity levels of DR and define the genes changed with DR progression. The landscape of immune cells and retinal cells in relation to DR progression has also been established by using a machine learning deconvolution algorithm, CIBERSORTx. For the first time, we identify a group of genes that are significantly increased in expression with the severity of DR. *CCND1* and *FCGR2B* among other severity-associated genes are key regulatory genes involved in inflammation during the DR progression. Deconvolution analysis reveals the significantly increased proportions of memory B cells, M2 macrophages and Müller glia with the severity of DR, while resting mast cells and naïve B cells are decreased in number with DR progression. These data provide detailed genetic and cellular characteristics of progressive DR.

## Methods

### Data accession

RNA-Seq raw counts were obtained from Gene Expression Omnibus (GSE160306, accessed on 28^th^ March 2022). Raw counts were converted to transcripts per million (TPM) in R (v4.1.0) for downstream analyses. Samples from human macular region were selected from the raw dataset using the subset function from the R package Seurat (v4.0.5).^5^ Detailed clinical information including patient gender, age and disease stages were accessed from GSE160310. Single-cell transcriptomes of human neural retina were directly downloaded from Human Cell Atlas (https://www.humancellatlas.org/, HCA, E-MTAB-7316, accessed on 15^th^ January 2022).^6^

### Identification of severity-associated genes in macular region from donors with DR at different stage

To identify genes related to the progressive severity of diabetic retinopathy, the selected RNA-Seq raw count was analysed by DESeq2 (v1.32.0),^7^ using the likelihood-ration test (LRT) as we previously described.^8^ Briefly, gene expression was determined by three variables, consisting of gender, age and disease status (severity). Significant severity-associated genes were determined by adjusted *p* < 0.05, controlled for gender and age. The resulting genes were then scaled to z-score and clustered using the degPatterns function from the R package DEGreport (v1.28.0). Gene clusters with consistent and progressive changes were selected for further analyses.

To further screen severity-associated genes, the TPM values of the genes from the selected cluster were analyzed by R package MASS (v7.3-54) and ordinal (v2019.12-10) using ordinal logistic regression (OLR) model.^9^ Using Cumulative Link Model function in ordinal,^9^ we adjusted gender and age to screen input genes that are associated with disease severity. Severity-associated genes were determined at *p* < 0.05. The resulting genes were represented in a forest plot and ranked according to its beta coefficient. A heatmap of severity-associated genes was generated by the pheatmap function from the R package NMF (v0.24.0).^10^

To assess gene-gene correlation of severity-associated genes, the raw counts were processed by Variance Stabilizing Transformation function in DESeq2,^7^ followed by statistical adjustment for gender and age using the Remove Batch Effect function in limma (v3.48.3).^11^ The correlation heatmap was plotted using R package corrplot (v0.92).^12^

### Functional enrichment analysis and gene expression visualization

Functional enrichment analyses (Gene Ontology Biological Processes) for significant severity-associated genes were performed with Metascape (https://metascape.org/gp/index.html#/main/step1),^13^ which provides more frequently updated bioinformatics analyses than DAVID.^14^

To visualize the expression of selected genes, the raw counts were processed by Variance Stabilizing Transformation function in DESeq2,^7^ followed by statistical adjustment for gender and age using the Remove Batch Effect function in limma (v3.48.3).^11^ Tukey boxplots was used to compare gene expression from patients with different DR status, with interquartile range boxes and 1.5x interquartile range whiskers. Two-tailed Mann-Whitney test was applied to assess pairwise comparisons in the plots, while Kruskal-Wallis test was used to perform statistical comparison across all groups of different disease status.

### Deconvolution analyses of the immune cells and retinal cells from donors with DR

The CIBERSORTx, a machine learning deconvolution algorithm that enables inference of cell-type-specific gene expression profiles,^15^ was applied to infer the estimated proportions of infiltrating immune cells or retinal cells of each bulk retinal transcriptome. We used the LM22 signature matrix to define 22 infiltrating immune cells^16^. Normalized data (TPM value) of the transcriptome were uploaded to the CIBERSORTx web portal (https://cibersortx.stanford.edu/), with the algorithm run using LM22 signature matrix at 100 permutations with B-mode batch correction.

To infer retinal cellular profile, we used a previously derived retinal signature matrix using the HCA scRNA-Seq dataset of the human retina.^8^ In brief, the retinal signature matrix contains rod, Müller glia, bipolar, cone, amacrine, microglia, ganglion, astrocytes, and horizontal cells. The retinal signature matrix was used to impute retinal cellular fraction from the bulk transcriptome at 100 permutations with S-mode batch correction. Statistical significance of cellular numbers associated with severity was assessed by an ordinal logistic regression model using the nonparametric Kruskal-Wallis test that allows for multi-factorial designs. The estimated coefficients and *p* value were presented as a forest plot with 95% confidence intervals. The estimated proportions of cell types were adjusted for gender and age to assess the effects by disease stages (severity) by extracting the residuals from fitting a generalized linear model with variables.

## Results

### Identification of severity-associated genes in the macula of patients with DR at different clinical stages

We analyzed a publicly available transcriptional dataset of the macular region from the retina of patients at different stages of DR and healthy controls. A total of 39 macular RNA-Seq profiles from controls (n=10) and cases from early to late DR stage (n=29) were analyzed (**Figure 1A**). The patient groups were previously classified according to the severity of DR progression, including diabetic (n=10), non-proliferative DR (NPDR, n=9), NPDR with DME (n=7) and PDR with DME (n=3).^17^ Of which, patients with NPDR with DME or PDR with DME were combined as one group of NPDR/PDR+DME (n=10) as the previous study described.^17^

**Figure 1.**
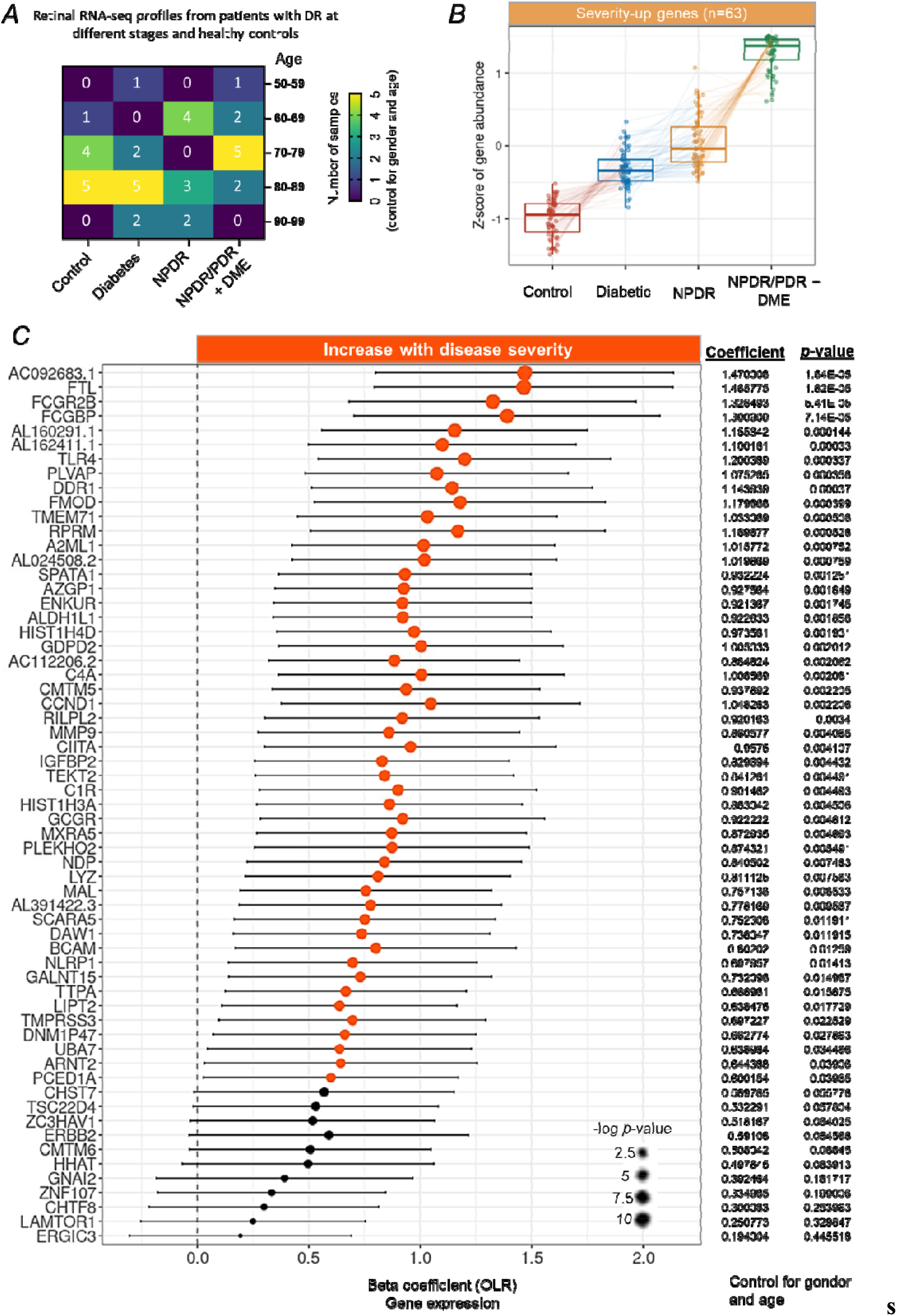
Identification of severity-associated genes in the macula from patient with degrees of DR. (A) Demographics of the human macular RNA-Seq profiles, detailed by age and different degrees of DR. (B) Tukey boxplots (interquartile range (IQR) boxes with 1.5× IQR whiskers) of severity-associated genes found in the macula of DR patients. Expression level of severity-up genes increases with the severity of DR (left, n=63). Severity-up genes were defined with DESeq2 two-sided likelihood-ratio test (adj. *p* < 0.05) by controlling for gender and age. Adjusted gene expressions were shown as z-score. (C) Forest plot of severity-associated genes significantly correlated with DR severity (p < 0.05, n=50). Positive coefficients indicate severity-associated genes increase in expression with DR severity. Statistical analyses of the correlation between the severity-associated genes and the DR progression were assessed by a nonparametric ordinal logistic regression model that controls for gender and age. Point sizes are scaled by statistical significance. Error bars represent 95% confidence intervals.

Gene expression levels in the retina were largely affected by age, gender and disease stages.^18^ By controlling for age and gender, we identified a cluster of genes (n=63) related to DR progression from the early to late stage (**Figure 1B**) using a likelihood-ratio test.^7^ Of note, the expression level of these genes were increased with the severity of DR. We therefore defined them as severity-up genes. To further screen genes significantly changed with the severity of DR, we assessed the correlation between severity-up genes and the severity of DR by using the OLR model that controls for gender and age. Our results identified 50 genes that are progressively increasing their expression level with the severity of DR (**Figure 1C**, hereafter referred to severity-associated genes, *p* <0.05). The remaining 13 severity-up genes were not significantly correlated to the severity of DR, thus excluding from the downstream analyses. A heatmap clearly demonstrated that the expression levels of 50 severity-associated genes were progressively increased with the severity of DR from the early (diabetic) to late stage (NPDR/PDR+DME), after adjustment for gender and sex (**Figure 2A**). Correlation analysis among these genes in terms of expression level revealed a cluster of genes (n=25) having the most interactions and strong correlations (|r| > 0.5) with other genes, suggesting their potentially important roles in the progression of DR (**Figure 2B**).

**Figure 2.**
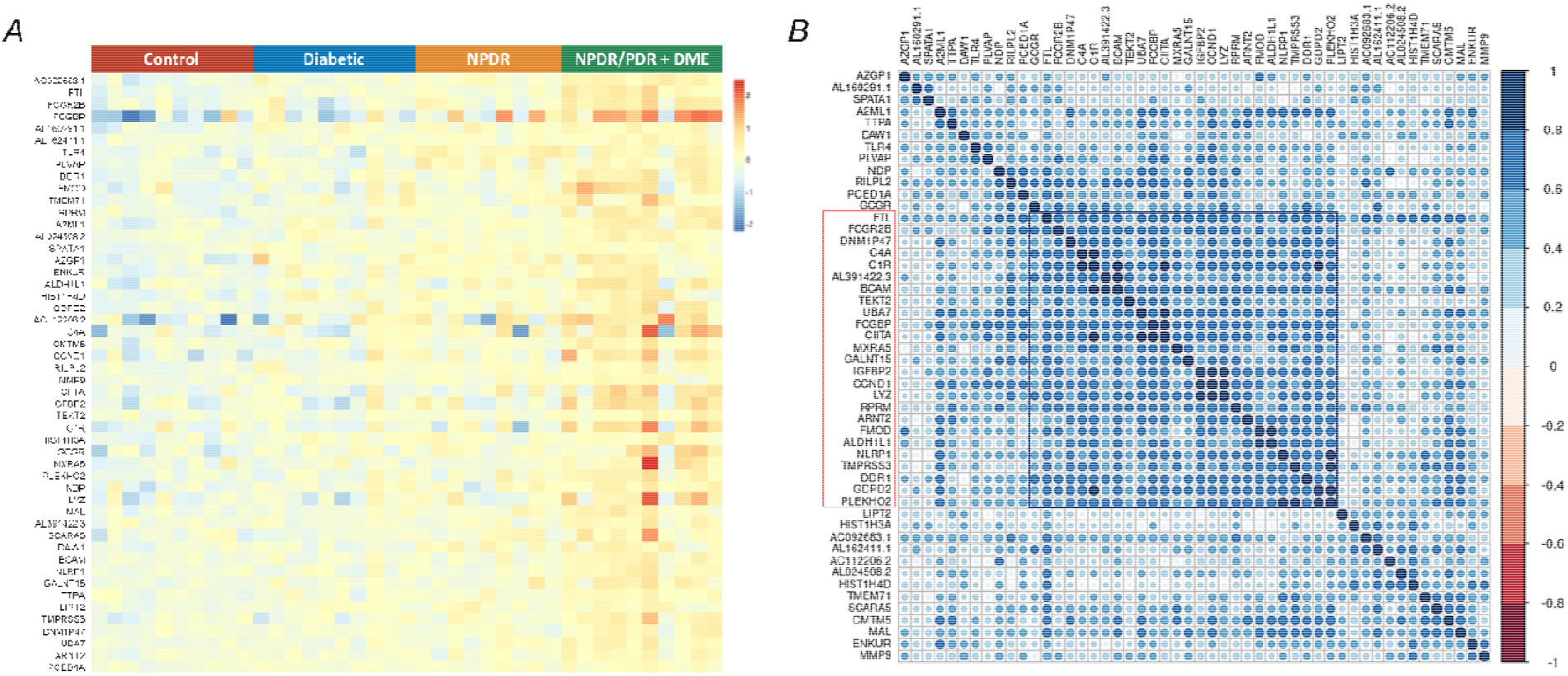
Expression profile and correlation analyses of severity-associated genes. (A) A heatmap of expression level of 50 severity-associated genes in all samples across different groups, adjusted for gender and age. (B) Heatmap of the correlation matrix across 50 severity-associated genes. Pearson’s correlation was calculated among the severity-associated genes to show the co-expression patterns of genes in the heatmap. Colour key denotes the Pearson’s correlation coefficient between genes. There are one clear cluster (highlighted in red and black) of severity-associated genes with strong positive correlation, adjusted by gender and age.

### Functional enrichment analyses of the severity-associated genes and identification of key genes involved in the progression of DR

To determine the functional roles of the severity-associated genes in the pathogenesis of DR, functional enrichment analysis (GO biological processes) was performed with Metascape. The results demonstrated significant enrichments mostly related to inflammation and immune response. Of which, negative regulation of cell differentiation, response to amyloid-beta, and inflammatory response rank top three among other GO terms (**Figure 3A**). Genes involved in the top three GO terms were combined to identify key genes that contribute to the progression of DR development, resulting in 7 genes with significant changes in expression with the severity of DR after controlling for gender and age (**Figure 3B**), including cyclin D1 (*CCND1*), Fc gamma receptor IIb (*FCGR2B*), matrix metallopeptidase 9 (*MMP9*), toll like receptor 4 (*TLR4*), alpha tocopherol transfer protein (*TTPA*), CKLF like MARVEL transmembrane domain containing 5 (*CMTM5*), and NLR family pyrin domain containing 1 (*NLRP1*). To further identify key severity-associated genes involved in the progression of DR, we overlapped the cluster of severity-associated genes (n=25) identified in **Figure 2B** with 7 genes identified in the functional enrichment analysis, leading to 3 genes shared by both clusters of genes (**Figure 3C**), including *CCND1, FCGR2B* and *NLRP1*. We focused on *CCND1* and *FCGR2B* and investigated their role in immune cells and retinal cells during DR. Given the expression pattern of *NLRP1* were not consistently increased with the severity of DR, we therefore excluded it for further analyses.

**Figure 3.**
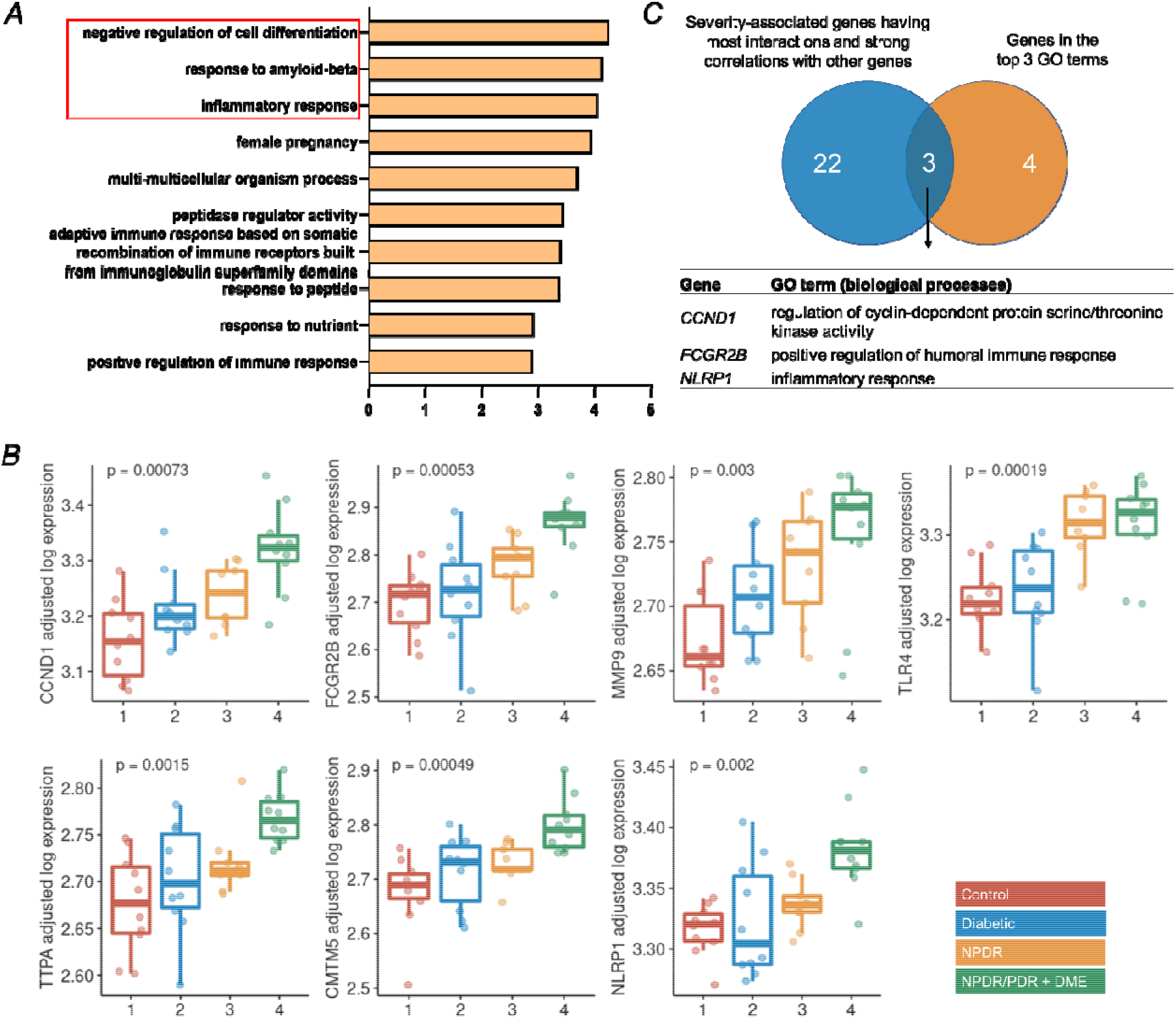
Functional enrichment analyses of severity-associated genes and identification of key genes involved in the DR progression. (A) Functional enrichment analyses (gene ontology annotations of biological processes) of 50 severity-associated genes by Metascape. The top 3 biological processes highlighted in red were selected for down-stream analysis. Genes with significant changes in these selected annotations include *CCND1, FCGR2B, MMP9, TLR4, TTPA, CMTN5* and *NLRP1*. (B) Tukey boxplots (interquartile range (IQR) boxes with 1.5× IQR whiskers) showing the expression of *CCND1, FCGR2B, MMP9, TLR4, TTPA, CMTN5* and *NLRP1*. Gene expression value are shown as log-transformed, controlled for gender and age. Statistical significance of difference was assessed by two-sided Kruskal-Wallis test on the adjusted expression values. (C) Overlapping the clustered genes from Figure 2B (n=25) and selected genes in Figure 3B (n=7) resulted in 3 genes, including *CCND1, FCGR2B* and *NLRP1*.

### The immune cellular landscape changed in the progression of DR

Given that severity-associated genes are strongly correlated to immune response, we next sought to understand the role of immune cells in the retina of patients with DR at different clinical stages. By using CIBERSORTx, we deconvoluted the macular transcriptomic data (**Supplementary Figure 1**) with the signature matrix of immune cells, LM22.^19^ We further assessed what immune cell types change in proportions with the severity of DR using an OLR model. By controlling for gender and age, we demonstrated that estimated proportions of memory B cells and M2 macrophages were significantly increased with the progression of DR, while the estimated proportions of resting mast cells and naïve B cells were significantly decreased with the progression of DR (**Figure 4A and Supplementary Figure 2**). Other immune cell types were neither found in the macular tissue nor their estimated proportions were not significantly altered with the progression of DR.

**Figure 4.**
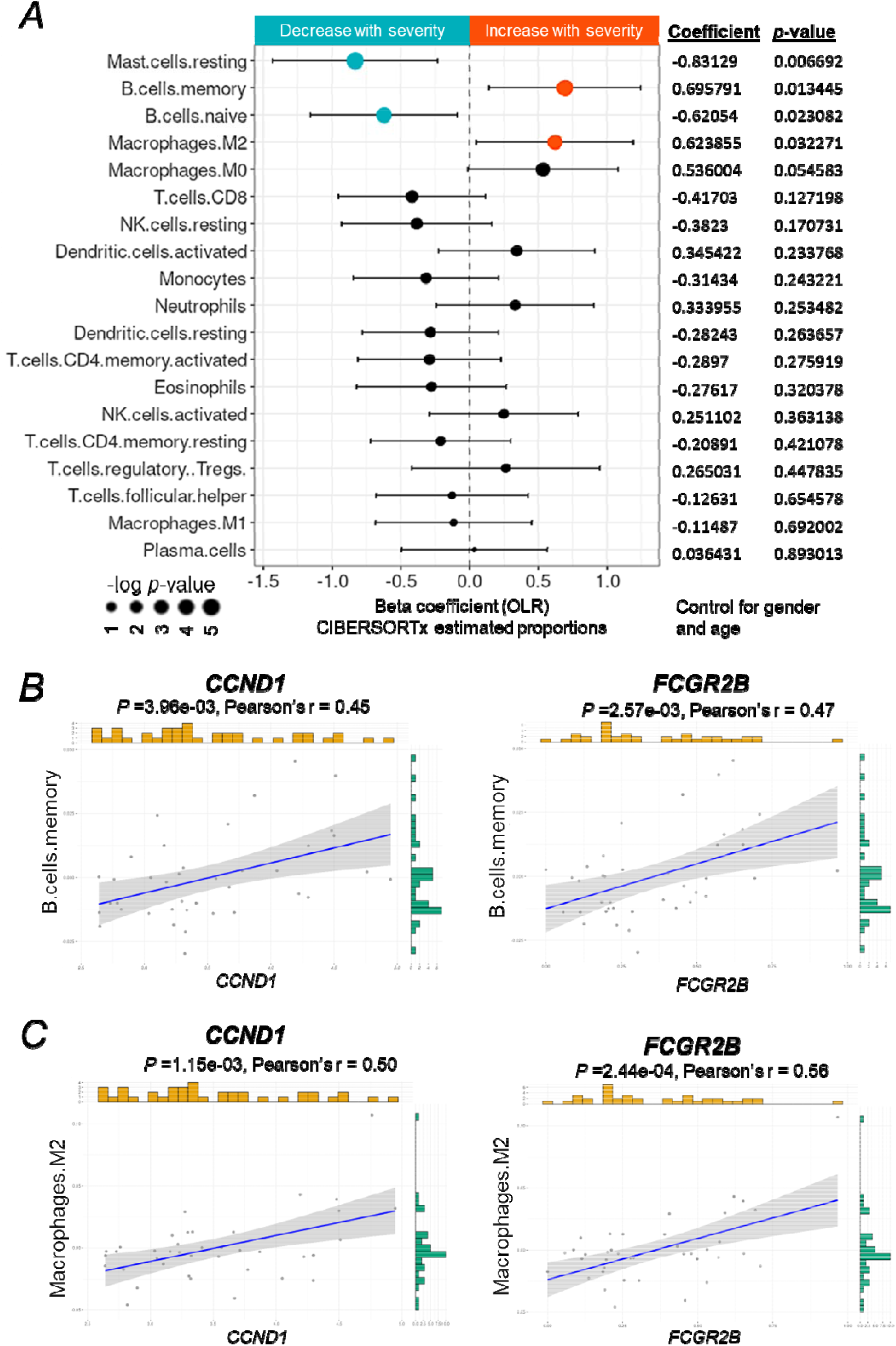
The immune cellular landscape of the macula of patients with DR and its relation to severity-associated genes. (A) Forest plot of estimated proportions of immune cells having significant correlation with the severity of DR (p < 0.05). Positive or negative coefficients indicate the proportion of immune cells increases or decrease with the severity of DR, respectively. Statistical analyses of correlation between the severity of DR and the proportions of immune cells were assessed by a nonparametric ordinal logistic regression model, adjusted for gender and age. Point sizes are scaled by statistical significance. Error bars represent 95% confidence intervals. (B and C) Analyses of the correlation between expression level of *CCND1* or *FCGR2B* and the estimated proportion of memory B cells or M2 macrophages, respectively, adjusted for gender and age. Pearson’s correlation coefficient was used to test the strength of linear relationships between gene expression and the estimated proportion of specific cell types. Grey dots denote individual samples (n=39). Blue lines denote regression lines.

We further studied the correlation between the expression level of two key genes identified above, *CCND1* and *FCGR2B*, and the proportions of the altered immune cell types, including memory B cells and M2 macrophages, using a Person’s regression analysis. Our results showed that expression level of both *CCND1* and *FCGR2B* have a moderate positive (0.3< r <0.7) correlation with the proportions of memory B cells (**Figure 4B**) and M2 macrophages (**Figure 4C**), respectively. These results suggest that *CCND1* and *FCGR2B* may be involved in the regulation of immune responses during the progression of DR mainly through M2 macrophages.

### The retinal cellular landscape changed in the progression of DR

A previous study demonstrated that the retinal structure particularly retinal nerve fiber layer is changed at the early onset of DR.^20^ However, quantitative changes in retinal cells during the progression of DR have not been reported. Here we deconvoluted the macular transcriptomes with CIBERSORTx to profile the estimated proportions of each of retinal cell types, using the scRNA-seq data from HCA as a reference (**Supplementary Figure 3**). We applied OLR model to control for gender and age and evaluated if the severity of DR affects the proportions of retinal cell types. The results revealed that the proportions of Müller glia were significantly increased with the progression of DR, while the proportions of other retinal cell type were altered but without significant difference (**Figure 5A and Supplementary Figure 4**). To confirm this finding, we further analyzed the expression profile of the severity-associated genes (40 genes were found in the scRNA-Seq dataset) in the human neural retina by using our previous scRNA-Seq data. Of the major retinal cell types, our results showed that most of the severity-associated genes were expressed in high percentage of Müller glia cells and astrocytes (**Figure 5B**). Given the estimated proportion of astrocytes are not significantly changed, we suspect Müller glia cells plays an important role in relation to the severity of DR.

**Figure 5.**
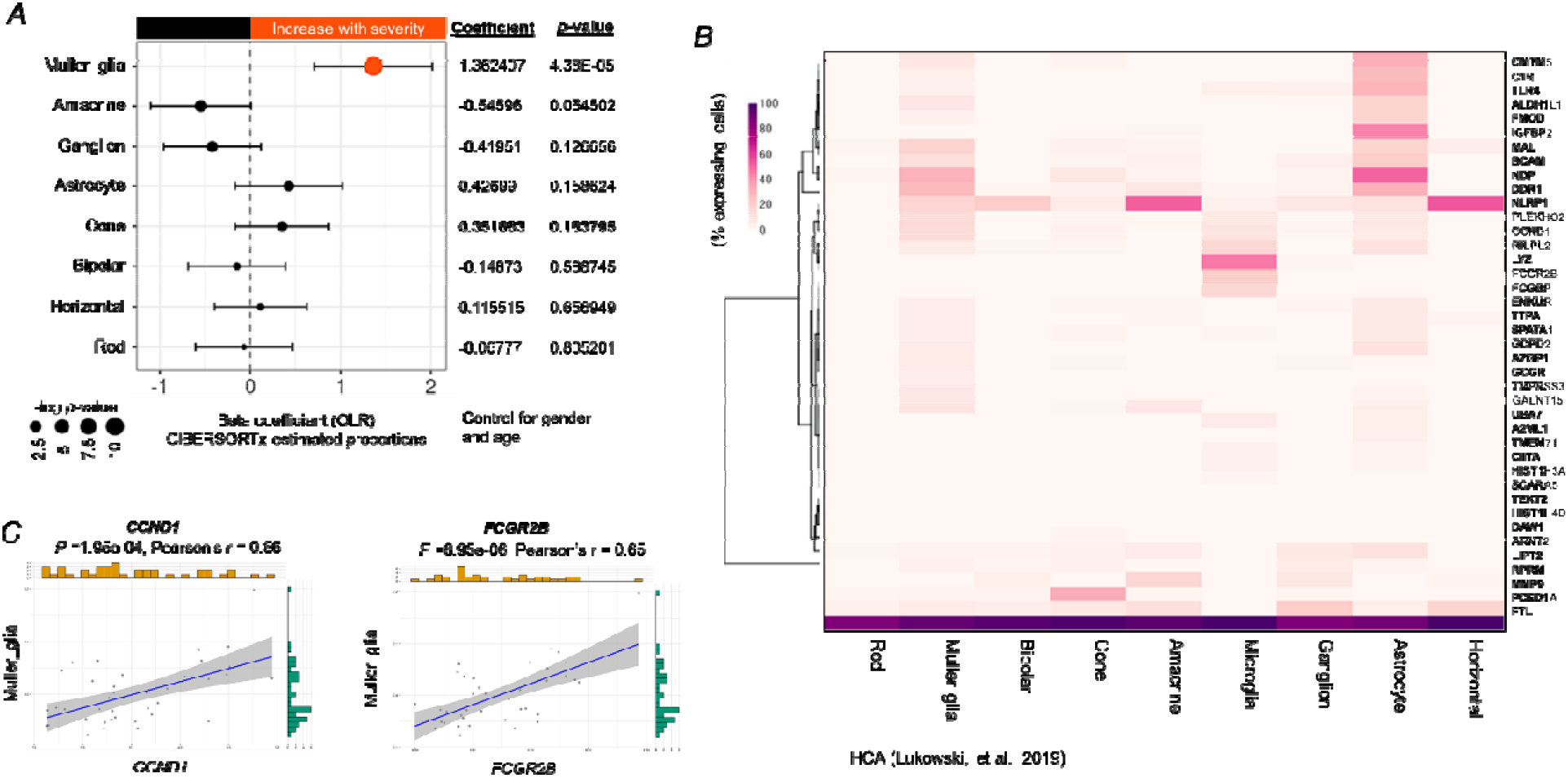
The cellular landscape of the macula of patients with DR and its relation to severity-associated genes. (A) Forest plot of estimated proportions of retinal cells having significant correlation with the severity of DR (p < 0.05). Positive or negative coefficients indicate the proportion of retinal cells increases or decrease with the severity of DR, respectively. Statistical analyses of correlation between the severity of DR and the proportions of retinal cells were assessed by a nonparametric ordinal logistic regression model, adjusted for gender and age. Point sizes are scaled by statistical significance. Error bars represent 95% confidence intervals. (B) Heatmap showing the percentage of retina cells expressing severity-associated genes (40 genes were found in the retinal scRNA-Seq dataset from the Human Cell Atlas), scaled by gene across the different cell types. (C) Analyses of the correlation between expression level of *CCND1* or *FCGR2B* and the estimated proportion of Müller glia, adjusted for gender and age. Pearson’s correlation coefficient was used to test the strength of linear relationships between gene expression and the estimated proportion of Müller glia. Grey dots denote individual samples (n=39). Blue lines denote regression lines.

Similarly, we investigated the relationship between the expression level of two key genes, *CCND1* and *FCGR2B*, and the proportion of Müller glia in DR. Our results showed that the expression level of *CCND1* and *FCGR2B* has a moderate positive (0.3< r <0.7) correlation with the proportion of Müller glia (**Figure 5C**), indicating that these genes may be modulated to promote increase of Müller glia with the severity of DR.

## Discussion

Genetic studies including linkage analyses, candidate gene association studies and genome-wide association studies have identified many risk loci or corresponding genes for DR.^3,21,22^ However, such studies tended to focus on one specific clinical stage of DR particularly the advanced stage, PDR and DME. Therefore, the resulting disease-causing genes from the genetic studies are not necessary to be associated with the progression of DR. Through application of LRT on transcriptome from patients with different degrees of DR along with OLR modelling, we identified 50 genes significantly increase in expression level from the early to late stage of DR (severity-associated genes) by controlling for gender and age. Among which, several severity-associated genes have been previously identified to be associated with DR in human or animal models, including *TLR4*,^23^ plasmalemma vesicle-associated protein (*PLVAP*),^24^ fibromodulin (*Fmod*),^25^ *C4A*,^26^ *MMP9*,^27^ insulin-like growth factor binding protein 2 (*IGFBP2*),^28^ and NLR Family Pyrin Domain Containing 1 (*NLRP1*),^29^ while none is reportedly increased with the progression of DR. Our results further validated those previous findings, indicating our transcriptomic analyses is reliable and comparable to other genetic studies. Although many severity-associated genes have not been reported in association with the progression of DR, interestingly, they are related to diabetes, such as alpha-2-Macroglobulin Like 1 (*A2ML1*),^30^ and *C1r*,^31^ suggesting their potential role in the pathogenesis of DR.

Inflammation is a non-specific response of immune system to harmful stimuli or stress that includes many functional and molecular mediators.^32^ Increasing evidence demonstrates that inflammation is crucial to the development of DR. Increased level of a variety of inflammatory cytokines and chemokines were observed in serum^33^ and ocular samples (aqueous and vitreous) from patients with PDR.^34^ However, it remains unclear how inflammation involves in the progression of DR and its relationship with the severity of DR due to the difficulty of sample collection. Our functional enrichment analyses revealed that the severity-associated genes are mainly involved in negative regulation of cell differentiation, inflammation and immune responses, suggesting these pathological processes are progressive during the development of DR.

We further identified cyclin D1 (*CCND1*) and Fc gamma receptor IIb (*FCGR2B*) are two key genes crucial to inflammation in DR. CCND1 protein is the critical gatekeeping protein regulating the transition from G1 phase to S phase via the restriction point of the cell cycle.^35^ Higher mRNA expression level of *CCND1* was actually observed in human diabetic islets by microarray and qRT-PCR.^36^ Heightened hepatic expression of *CCND1* was also found in animal models of obesity or diabetes due to hyperinsulinemia,^37^ suggesting a close relationship between *CCND1* and diabetes. The study with the macular transcriptomic data from DR patients that we analyzed in the present study also found *CCND1* was one of genes identified with the strongest significant expression changes.^17^ Our correlation analysis among severity-associated genes confirmed that *CCND1* has a strong interaction with most of other severity-associated genes, indicating its critical role in mediating or regulating different genes that together contributes to the progression of DR. *FCGR2B* protein is a low affinity receptor for the Fc region of immunoglobulin gamma complexes.^38^ *FCGR2B* involves in many aspects of inflammatory and immune responses as well as in the complex regulation of defence against infection.^39^ However, the regulation of *FCGR2B* expression is complicated depending on the cell types. For example, interferon gamma, a signature proinflammatory cytokine, escalates *FCGR2B* mRNA and cell surface expression by lipopolysaccharide-stimulated B cells while reduces it on monocytes, a process keeping immune response away from antibody production to pathogen clearance.^39^ Increase levels of *FCGR2B* has been observed in the pancreata of autoantibody-positive at-risk individuals with type 1 diabetes versus controls,^40^ and in the retina of a rat model with type 1 diabetes induced by streptocotocin.^31^ Similar to *CCND1, FCGR2B is* also one of severity-associated genes with strong interaction with other genes. Altogether, our results are in line with previous findings and further confirmed both *CCND1* and *FCGR2B* increase in mRNA expression with the severity of DR, suggesting their potential role as biomarkers to monitor the progression of DR.

In the early stages of DR when blood retinal barrier is intact, the immune system is mildly activate and retinal microglia plays a major role in the para-inflammatory response, an important process to maintain retinal homeostasis.^41^ Consistent stimulation of damage-associated molecules induced by diabetes in the eye cause maladaptation of the innate immune systems, ultimately promoting the development of DR.^41^ Circulating immune cells penetrate to the retina and result in chronic inflammation, leading to retina vascular and neuronal damage in the late stage.^42^ Despite the importance of immune system in the pathogenesis of DR, few studies have investigated the changes of immune cell types over the progression of DR. Using the deconvolution method with CIBERSORTx, we systemically analyzed immune cellular profiles containing 22 immune cells and identified significant changes in proportions of resting mast cells, memory B cells, naïve B cells and M2 macrophages in the macular region from donors with different degrees of DR. Mast cells are tissue resident with heterogenous phenotypes tuned by inflammatory stimui.^43^ We suspect that inflammation in DR results in the phenotype change of mast cells from resting to activated, for which proportions of resting mast cells are significantly decreased over severity of DR. Memory B cells are the populations of cells provide long-term humoral immunity.^44^ A study revealed the chronic inflammation triggers increase of activated memory B cells in patients with erythema nodosum leprosum, an inflammatory complication of leprosy.^45^ Given DR is a progressive disease featured with chronic inflammation, it is reasonable to see increased and decreased proportions of memory B cells and naïve B cells, respectively with the severity of DR. Macrophages are heterogeneous and featured with various function once activated.^46^ Our previous study demonstrated the significant increase of M2 macrophages in the retina of the rat model of retinal neovascularization and of patient with proliferative diabetic retinopathy by deconvolution analysis.^47^ Our results confirmed previous findings that the proportions of M2 macrophages are increased with the severity of DR. Correlation analyses showed *CCND1* and *FCGR2B* have a moderate and moderate positive correlation with memory B cells and M2 macrophages, respectively, suggesting both genes may be crucial in regulating activation of M2 macrophages.

Despite studies have shown morphological or structural changes in retina cells from patients with DR, such as Müller glia and microglia cells,^48,49^ few assessed changes in proportions of each of retinal cells from patients with DR. Using CIBERSORTx, we for the first time quantified the proportions of all retinal cells in the macular region from patients with different degrees of DR. We revealed that the proportion of Müller glia is significantly increased with the severity of DR, while changes of other retinal cells were observed but without significant difference. Expression of *CCND1* and *FCGR2B* is moderate positive related to the proportions of Müller glia, indicating their importance in activation of Müller glia. Müller glia is important to maintain retinal function and heath because it spans the entire width of the retina and contact to every other cell type. It is believed that Müller glia become activated in diabetic retinopathy, characterized by the increased expression of glial fibrillary acidic protein, a marker of reactive gliosis.^49,50^ However, some histological studies also reported Müller cells die during the DR progress possibly due to the increased caspase-1 activity and IL-1β production following exposure to hyperglycemic conditions.^50,51^ More studies are required to determine the mechanisms of Müller glia activation and death to pinpoint whether all Müller glia are equally affected by hyperglycemia.

### Limitations

There were several limitations of this study. First, the sample size in each group of DR stage is relatively small, which likely affects the statistical power to identify high number of severity-associated genes defined by LRT. In addition, the current study is based on in silico analysis transcriptomic data. Future biological studies that investigate the functionality of *CCND1* and *FCGR2B* in memory B cells and M2 macrophages as well as Müller glia would be important to understand their role in progressive DR.

## Conclusions

The findings reveal a group of genes particularly *CCND1* and *FCGR2B* significantly increase in expression with the severity of DR, controlling for other factors affecting gene expression, such as gender and age. Moreover, proportions of memory B cells, M2 macrophages and Müller glia are significantly increased with the severity of DR. Our analyses provide potential avenues for subsequent research to study the mechanisms of DR progression.

## Data Availability

Analysis scripts will be made available upon request. RNA-Seq raw counts were obtained from Gene Expression Omnibus (GSE160306, accessed on 28th March 2022).

## Code availability

Analysis scripts will be made available upon request.

## Acknowledgements

We thank Ryan D. Chow for sharing codes for data analyses at Github.

## Author contribution

J-H.W. and G-S.L. conceived and designed the study. J-H.W. developed the analysis approach, performed all data analyses, and generated figures. J-H.W., R.C.B.W., and G-S.L. prepared the manuscript. G-S.L. supervised the work.

